# Toward Clinical Implementation of Polygenic Scores for Substance Use Disorders: A Multi-Ancestry Study

**DOI:** 10.64898/2026.07.03.26357210

**Authors:** Dongbing Lai, Michael Zhang, Tae-Hwi Schwantes-An, Marcus R. Breese, Karen Chartier, Christina M. Sheerin, Martin H. Plawecki, Changyong Guo, Yao-Ying Ma, Zhiping P. Pang, Howard J. Edenberg, Tatiana Foroud, Yunlong Liu

## Abstract

**Objective:** To develop and validate clinically relevant polygenic scores (PGS) for alcohol (AUD), cannabis (CanUD), opioid (OUD), tobacco (TUD), and polysubstance use disorders (polySUD) across African (AA), European (EA), and Latinx (LA) ancestry populations.

**Methods:** Using multiple genome-wide association study summary statistics and PGS methods, substance use disorder PGS were developed and evaluated in Indiana Biobank samples (IB, N: 1,356-24,989), then top-performing PGS were validated in All of Us Research Program samples (AOU, N: 62,389-209,952). Case and controls were defined using ICD-9/10 codes. All participants were aged 18 years or older (>=21 years for AUD controls). Clinical relevance was defined as an odds ratio (OR) >=2 for individuals with the highest PGS determined based on disorder prevalence compared to everyone else.

**Results:** In EA and LA, all PGS achieved clinically relevant performance in both IB and AOU (ORs: 2.00-9.10; P <= 3.87E-4). In AA, PGS met this threshold in IB (ORs: 2.02-2.71; P <= 2.20E-4) but not in AOU (ORs: 1.28-1.56; P <=0.03). Overall, OUD PGS showed the strongest associations in most analyses, followed by CanUD and polySUD. Generally, compared to female PGS, male PGS had higher or comparable ORs, but the differences were not significant except AUD PGS in AOU LA.

**Conclusions:** PGS demonstrated clinically meaningful risk prediction for substance use disorders in EA and LA, supporting the feasibility of future clinical implementation for population-level screening. However, reduced performance in AA underscores the urgent need for more genetic studies in that population.

## INTRODUCTION

Substance use disorders (SUDs) are chronic and relapsing conditions that have devastating consequences on individuals, their families, and society. SUDs are highly prevalent. In 2022, 17.3% of individuals in the U.S. met criteria for at least one SUD within the past 12 months, including 8.7% of adolescents aged 12 to 17 years(1). Additionally, SUDs significantly increase the risks for other diseases and mortality(2, 3), and for those younger than 45, drug overdose is the major cause of death(4).

SUDs are preventable, and prevention is most effective when high-risk individuals are identified early and targeted prevention programs are implemented promptly(5, 6). However, commonly used screening tools (e.g. Alcohol, Smoking, and Substance Involvement Screening Test(7)) are designed primarily for intervention because they rely on information about risky substance use. For many individuals, this means that they may already progress beyond the point that prevention programs can be effective. Importantly, for adolescents and young adults, substance use can adversely affect neurodevelopment. While family history is widely used to predict risk without relying on substance use information, it can be unavailable or incomplete. Furthermore, studies have demonstrated that SUDs are polygenic disorders(8–14); therefore, many individuals with SUDs are not expected to have a positive family history based on the polygenic theory(15–17). Complementary tools are needed to improve risk prediction beyond family history.

The estimated heritability of each SUD is ∼50%(18), supporting the use of polygenic scores (PGS) as a risk prediction tool. PGS aggregate the effects of risk-associated single nucleotide polymorphisms (SNPs) and can be assessed before initiation of substance use, potentially enabling earlier and more effective prevention strategies. SNPs and their effect sizes were derived from genome-wide association studies (GWAS; referred to as the discovery datasets), whereas PGS were developed and tested in independent datasets (referred to as the target datasets). To achieve clinical relevance, individuals with the highest PGS should exhibit an odds ratio (OR) ≥2 for developing SUD compared to everyone else in the same population—comparable to the predictive value of family history, pathogenic variants, and risk factors for disorders such as cardiovascular disease, diabetes, breast and prostate cancers that are currently used in medical practice(19, 20). However, several challenges limit the development of PGS with sufficient predictive performance and generalizability for clinical implementation. First, discovery and target datasets often differ substantially in ascertainment strategies, diagnostic criteria, and participant characteristics. Consequently, relying on a single discovery GWAS may fail to identify the PGS with optimal predictive performance. This challenge is particularly pronounced for underrepresented populations, such as individuals of African (AA) and Latinx (LA) ancestry, because large-scale European ancestry (EA) GWAS are frequently leveraged to improve statistical power, yet multiple strategies exist for leveraging these EA GWAS. Second, substantial heterogeneity across target datasets can limit the generalizability of PGS developed in a single cohort. Third, different PGS methods rely on different assumptions, and no single method consistently performs best across traits and datasets. Therefore, restricting analyses to one or a few methods may overlook more predictive PGS.

To address these challenges, Khera and colleagues developed a two-stage optimization and validation framework and developed the first PGS being tested in clinical trials (20–22). The two-stage framework systematically evaluated multiple GWAS and PGS methods to identify the optimal one in the screening stage, then independently validated them in the testing stage to improve both predictive power and generalizability of PGS. Building on this framework, we sought to develop clinically relevant and generalizable PGS for the five most prevalent SUDs: AUD, cannabis (CanUD), opioid (OUD), tobacco (TUD), and polysubstance (polySUD) use disorders across AA, EA, and LA populations. Two large biobank samples, Indiana Biobank (IB) (23, 24) and All of Us Research Program (AOU, version 8) (25) were used in the screening stage and testing stage, respectively.

## METHODS

### Study overview

The study design is shown in **Figure 1**. For each SUD, the largest GWAS to date was used as the discovery dataset. Additionally, the largest cross SUD (crossSUD) GWAS(26), which searched for SUD-shared SNPs, was also used. Additionally for AA and LA, different ways of leveraging EA GWAS were used. IB and AOU were used as the target datasets, and we used IB at the screening stage due to its higher SUD prevalences. At the screening stage, pruning and thresholding (P+T)(27, 28), and two state-of-the-art Bayesian-based PGS methods, PRS-CS(29) and PRS-CSx(30) (for AA and LA only), were used. For each SUD in each ancestry, to avoid IB-specific findings and increase PGS generalizability, we selected the 10 best PGS based on ORs. These PGS were then validated in AOU, and the one with the highest OR was selected as the final PGS. Statistical significance in the testing stage was defined using a Bonferroni-corrected threshold of P-value < 0.005.

**Figure 1:**
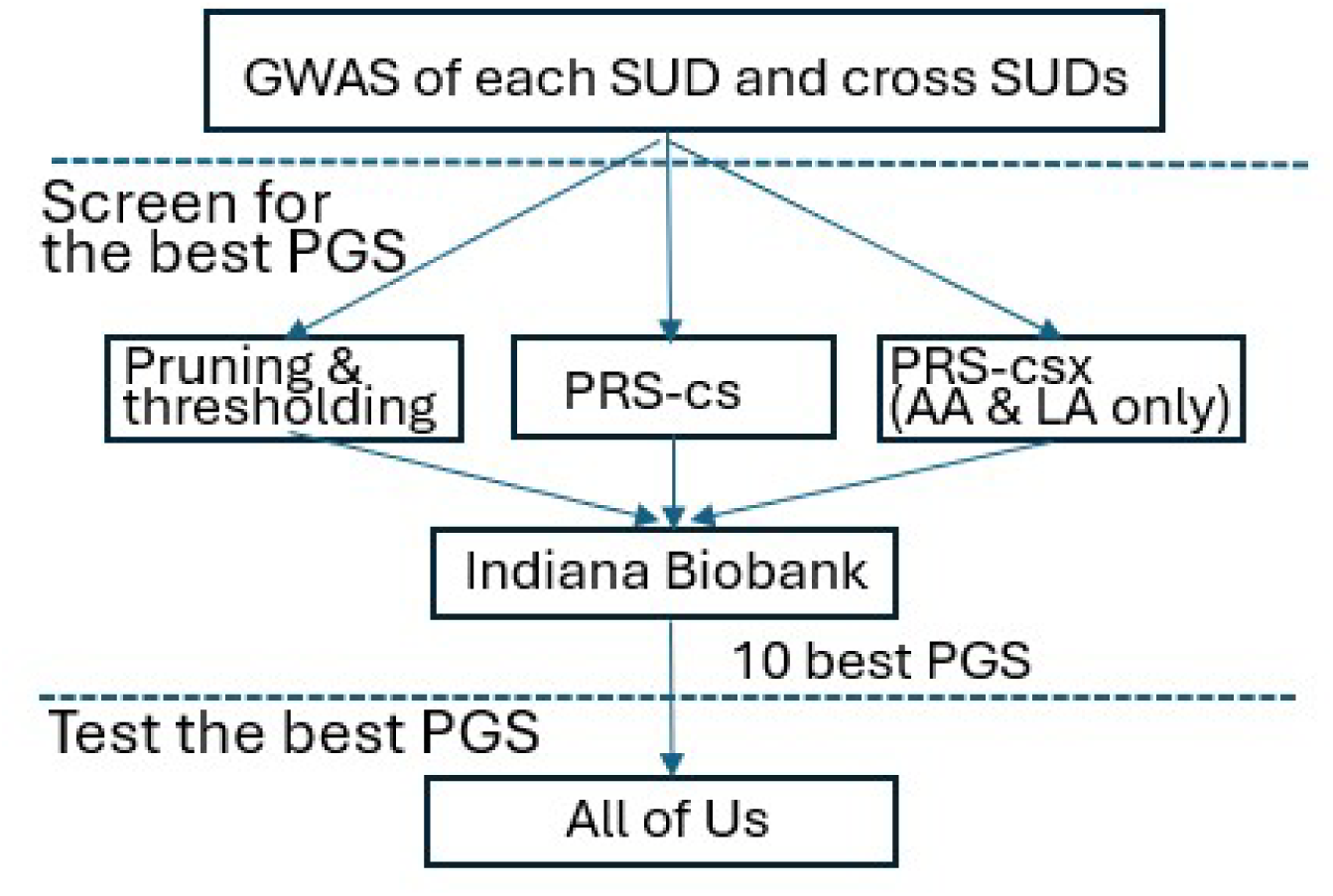
workflow of the study.

This study was approved by the institutional review board of Indiana University School of Medicine. All participants provided written consent.

### Discovery datasets and PGS methods

*<u>AUD GWAS:</u>* AA and EA GWAS of problematic alcohol use(9) were downloaded from https://medicine.yale.edu/lab/gelernter/stats/. LA GWAS of AUD(9) was obtained from the database of genotypes and phenotypes (dbGaP, phs001672). Three large study cohorts were included in EA GWAS: the Million Veteran Program(9), the UK Biobank(8), and the FinnGen consortium(31); as in our previous studies(32, 33), we only retained SNPs that had concordant effects to exclude false positives and cohort-specific findings. Three AA and three LA GWAS were used: 1) AA or LA only GWAS; 2) cross-ancestry meta-analysis of AA and EA or LA and EA, noting many disease-associated SNPs are shared among different ancestries; 3) cross-ancestry meta-analysis but only retaining SNPs having concordant effects in different populations (population-concordant SNPs) combined with AA- or LA-specific SNPs, as SNPs with discordant effects across ancestries may decrease PGS predictive performance(23, 26, 34).

*<u>CanUD GWAS:</u>* AA and EA CanUD GWAS(10) were downloaded from https://medicine.yale.edu/lab/gelernter/stats/. LA CanUD GWAS(10) was not available yet and thus we used EA CanUD GWAS. For AA, as in AUD, we also used three GWAS: AA only, cross-ancestry meta-analysis of AA and EA, cross-ancestry meta-analysis of AA and EA but retaining population-concordant and AA-specific SNPs.

*<u>OUD GWAS:</u>* AA, EA, and LA OUD GWAS(11) were obtained from dbGaP (phs001672). For AA and LA, three GWAS were used: AA or LA only, cross-ancestry meta-analysis of AA and EA or LA and EA, cross-ancestry meta-analysis of AA and EA or LA and EA but retaining population-concordant and AA-specific or LA-specific SNPs.

*<u>TUD GWAS:</u>* AA and EA TUD GWAS were obtained from the corresponding author of the largest TUD GWAS(14). LA TUD GWAS(14) was not available yet and thus we used EA TUD GWAS. For AA, three GWAS were used: AA only, cross-ancestry meta-analysis of AA and EA, cross-ancestry meta-analysis of AA and EA but retaining population-concordant and AA-specific SNPs.

*<u>crossSUD GWAS:</u>* These are all from our previous work(26). For AA and LA, three GWAS were used: AA or LA only, cross-ancestry meta-analysis of AA and EA or LA and EA, cross-ancestry meta-analysis of AA and EA or LA and EA but retaining population-concordant and AA-specific or LA-specific SNPs. Note, while crossSUD was mainly for developing PGS for polySUD, it was also included in constructing PGS for AUD, CanUD, OUD, and TUD because the identified SNPs were SUD-shared.

For P+T, we varied P-value thresholds (1, 0.5, 0.2, 0.1, 0.05, 0.01, 0.005, 0.001, 0.0005, 0.0001, 0.00005, 0.00001, 0.000005, 0.000001, 0.0000005, 0.0000001, 0.00000005), linkage disequilibrium (LD) r^2^ (0.1, 0.2, 0.3, 0.4, 0.5), and physical distance to calculate r^2^ (250kb and 500kb), resulting in 170 sets of SNPs. As Bayesian based methods are unstable, and different analyses generates different results; therefore, we repeated PRS-CS and PRS-CSx 10 times to identify the one with the highest OR. For all three PGS methods, LD was determined by using the 1000 Genomes Project AA, EA, and LA samples(35).

### Target datasets

IB is a state-wide collaboration that provides centralized processing and storage of specimens that are linked to participants’ electronic health records from Indiana University School of Medicine health care systems(23, 24). AOU is a national research resource aimed at recruiting participants with diverse conditions from diverse backgrounds(25). For both IB and AOU, SUD status was determined based on ICD 9/10 codes. All participants were ≥18 years old except AOU controls, which were ≥21 years old. Detailed information regarding data processing and definition of cases and controls are in **Supplemental Methods**.

### PGS calculation and statistical analysis

PGS were calculated using PLINK(36, 37), and we refer to these PGS as the raw PGS. Since genetic ancestries, even those fine scale ones in EA, have substantial impact on PGS distributions(19, 38), we calculated PGS residuals (resPGS) to make PGS independent of genetic ancestries. First, raw PGS were regressed on the first 10 principal components (PCs) of genetic ancestry using linear regressions. Note, this was performed using control samples only because PCs may capture genetic differences due to disease status and thus including cases can generate biased estimates. Then the estimated coefficients from the linear regressions were used to calculate the predicted PGS for all samples, and resPGS were calculated as predicted PGS minus raw PGS. All individuals shared the same resPGS distribution as they were adjusted for the effects of genetic ancestry.

To determine the resPGS thresholds for defining high-risk groups, we performed a grid-search to systematically evaluate candidate percentile cutoffs. Percentages of those at high-risk should not be larger than SUD prevalences to avoid overclassification of individuals unlikely to develop SUDs (but could be smaller as some people could develop SUDs due to non-genetic factors). Therefore, we started with 2.5% of individuals with the highest PGS and incrementally increased the threshold by 2.5% until it approached—but did not exceed—the SUD prevalences. If the prevalence was <2.5%, then the prevalence was used. We compared those with the highest resPGS to everyone else using logistic regression with sex as a covariate. The threshold for the best PGS was determined if the OR ≥2 and identified the largest number of high-risk individuals (i.e., the percentage was the closest one to the prevalence). For the best PGS, we also performed sex-stratified analyses given sex differences in SUD prevalences.

## RESULTS

The characteristics of IB and AOU cohorts are summarized in **Table 1**. In both datasets, the sample sizes were largest for EA, followed by AA, then LA; and females comprised more than half of participants (57%-66%). TUD had the highest prevalences (9.84%-36.84%), followed by polySUD (4.47%-18.53%), AUD (4.46%-14.02%), OUD (2.46%-7.31%), and CanUD (1.42%-8.41%). For all SUDs, across both cohorts, AA had the highest prevalences. EA had higher prevalences than LA in IB, whereas prevalences were comparable in AOU. Except OUD in IB EA and IB LA, SUDs prevalences were higher in males than in females.

**Table 1:** Summary of the target datasets.

| Ancestry | Cohort | Phenotypes | All samples |  |  |  | Male |  |  |  | Female |  |  |  |
| --- | --- | --- | --- | --- | --- | --- | --- | --- | --- | --- | --- | --- | --- | --- |
|  |  |  | # Case | # Control | # Total | Prevalence | # Case | # Control | # Total | Prevalence | # Case | # Control | # Total | Prevalence |
| AA | IB | AUD | 534 | 3,274 | 3,808 | 14.02% | 305 | 1,004 | 1,309 | 23.30% | 229 | 2,270 | 2,499 | 9.16% |
|  | IB | CanUD | 321 | 3,498 | 3,819 | 8.41% | 148 | 1,165 | 1,313 | 11.27% | 173 | 2,333 | 2,506 | 6.90% |
|  | IB | OUD | 278 | 3,524 | 3,802 | 7.31% | 104 | 1,184 | 1,288 | 8.07% | 174 | 2,340 | 2,514 | 6.92% |
|  | IB | TUD | 1,406 | 2,411 | 3,817 | 36.84% | 586 | 725 | 1,311 | 44.70% | 820 | 1,686 | 2,506 | 32.72% |
|  | IB | polySUD | 706 | 3,105 | 3,811 | 18.53% | 364 | 945 | 1,309 | 27.81% | 342 | 2,160 | 2,502 | 13.67% |
|  | AOU | AUD | 5,254 | 63,719 | 68,973 | 7.62% | 3,327 | 26,197 | 29,524 | 11.27% | 1,927 | 37,522 | 39,449 | 4.88% |
|  | AOU | CanUD | 3,100 | 65,997 | 69,097 | 4.49% | 1,742 | 27,817 | 29,559 | 5.89% | 1,358 | 38,180 | 39,538 | 3.43% |
|  | AOU | OUD | 2,627 | 66,493 | 69,120 | 3.80% | 1,520 | 28,047 | 29,567 | 5.14% | 1,107 | 38,446 | 39,553 | 2.80% |
|  | AOU | TUD | 13,630 | 55,754 | 69,384 | 19.64% | 6,842 | 22,840 | 29,682 | 23.05% | 6,788 | 32,914 | 39,702 | 17.10% |
|  | AOU | polySUD | 7,128 | 62,143 | 69,271 | 10.29% | 4,245 | 25,394 | 29,639 | 14.32% | 2,883 | 36,749 | 39,632 | 7.27% |
| EA | IB | AUD | 2,467 | 22,382 | 24,849 | 9.93% | 1,399 | 8,404 | 9,803 | 14.27% | 1,068 | 13,978 | 15,046 | 7.10% |
|  | IB | CanUD | 711 | 24,246 | 24,957 | 2.85% | 365 | 9,504 | 9,869 | 3.70% | 346 | 14,742 | 15,088 | 2.29% |
|  | IB | OUD | 1,335 | 23,654 | 24,989 | 5.34% | 509 | 9,331 | 9,840 | 5.17% | 826 | 14,323 | 15,149 | 5.45% |
|  | IB | TUD | 6,324 | 18,607 | 24,931 | 25.37% | 2,950 | 6,914 | 9,864 | 29.91% | 3,374 | 11,693 | 15,067 | 22.39% |
|  | IB | polySUD | 2,429 | 22,479 | 24,908 | 9.75% | 1,256 | 8,598 | 9,854 | 12.75% | 1,173 | 13,881 | 15,054 | 7.79% |
|  | AOU | AUD | 9,334 | 200,152 | 209,486 | 4.46% | 5,854 | 78,138 | 83,992 | 6.97% | 3,480 | 122,014 | 125,494 | 2.77% |
|  | AOU | CanUD | 2,986 | 206,866 | 209,852 | 1.42% | 1,725 | 82,355 | 84,080 | 2.05% | 1,261 | 124,511 | 125,772 | 1.00% |
|  | AOU | OUD | 5,344 | 204,560 | 209,904 | 2.55% | 2,544 | 81,558 | 84,102 | 3.02% | 2,800 | 123,002 | 125,802 | 2.23% |
|  | AOU | TUD | 22,798 | 187,154 | 209,952 | 10.86% | 10,704 | 73,436 | 84,140 | 12.72% | 12,094 | 113,718 | 125,812 | 9.61% |
|  | AOU | polySUD | 9,382 | 200,561 | 209,943 | 4.47% | 5,300 | 78,818 | 84,118 | 6.30% | 4,082 | 121,743 | 125,825 | 3.24% |
| LA | IB | AUD | 102 | 1,254 | 1,356 | 7.52% | 54 | 500 | 554 | 9.75% | 48 | 754 | 802 | 5.99% |
|  | IB | CanUD | 36 | 1,342 | 1,378 | 2.61% | 17 | 546 | 563 | 3.02% | 19 | 796 | 815 | 2.33% |
|  | IB | OUD | 48 | 1,313 | 1,361 | 3.53% | 15 | 536 | 551 | 2.72% | 33 | 777 | 810 | 4.07% |
|  | IB | TUD | 246 | 1,133 | 1,379 | 17.84% | 122 | 443 | 565 | 21.59% | 124 | 690 | 814 | 15.23% |
|  | IB | polySUD | 106 | 1,268 | 1,374 | 7.71% | 53 | 509 | 562 | 9.43% | 53 | 759 | 812 | 6.53% |
|  | AOU | AUD | 2,872 | 59,517 | 62,389 | 4.60% | 1,925 | 19,953 | 21,878 | 8.80% | 947 | 39,564 | 40,511 | 2.34% |
|  | AOU | CanUD | 1,228 | 61,601 | 62,829 | 1.95% | 723 | 21,277 | 22,000 | 3.29% | 505 | 40,324 | 40,829 | 1.24% |
|  | AOU | OD | 1,546 | 61,330 | 62,876 | 2.46% | 913 | 21,111 | 22,024 | 4.15% | 633 | 40,219 | 40,852 | 1.55% |
|  | AOU | TUD | 6,195 | 56,774 | 62,969 | 9.84% | 3,246 | 18,812 | 22,058 | 14.72% | 2,949 | 37,962 | 40,911 | 7.21% |
|  | AOU | polySUD | 3,265 | 59,679 | 62,944 | 5.19% | 2,051 | 20,002 | 22,053 | 9.30% | 1,214 | 39,677 | 40,891 | 2.97% |

Note: AA: African ancestry; EA: European ancestry; LA: Latinx ancestry; IB: Indiana Biobank; AOU: All of Us research program; AUD: alcohol use disorder; CanUD: cannabis use disorder; OD: opioid use disorder; TUD: tobacco use disorder; polySUD: poly substance use disorder.

The top 10 best PGS for each SUD in each ancestry are listed in **Supplemental Tables 1-15**. The best-performing PGS are summarized in **Figure 2** and **Supplemental Table 16**. In EA and LA, ORs in IB ranged from 2.00 to 9.10 (P ≤3.87×10), ORs in AOU ranged from 2.23 to 3.82 (P ≤ 9.13×10 ²²), and half of PGS having ORs ≥3.21. Between EA and LA, ORs were comparable in AOU but were higher in LA than in EA in IB (with larger 95% confidence intervals (CIs) due to the smaller sample sizes). In AA, all PGS except for OUD were significant (P ≤2.71×10), but none achieved the predefined clinical relevance threshold in AOU. Overall, CanUD, OUD, and polySUD PGS had higher ORs. In EA and LA, the thresholds used to define high-risk groups ranged from 1.42% to 17.5%. Except for AUD and OUD in IB, all thresholds exceeded 50% of the corresponding disorder population prevalences, and half were greater than 90% of the prevalences, indicating large numbers of individuals at high-risk can be identified.

**Figure 2:**
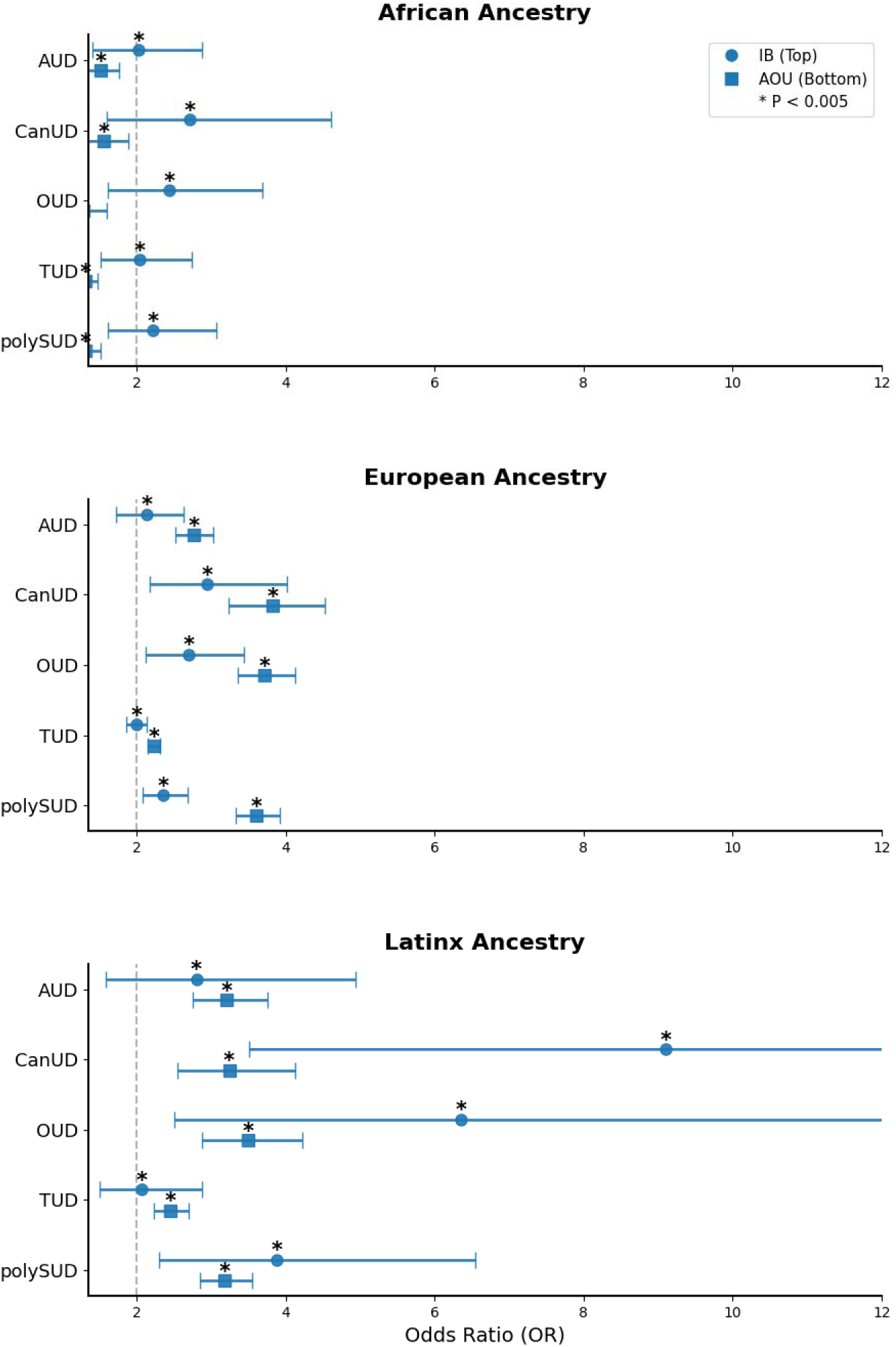
The best PGS in IB and AOU. Note: AUD: alcohol use disorder; CanUD: cannabis use disorder; OUD: opioid use disorder; TUD: tobacco use disorder; polySUD: poly substance use disorder; Dashed line indicates OR=2. To improve readability, the x-axis was limited to 12. The 95% confidence intervals for IB CUD and OUD extend beyond the displayed range and are truncated.

For the discovery datasets, except for AUD, CanUD, and TUD in AA, all best-performing PGS were derived from crossSUD GWAS (**Supplemental Table 16**). In AA and LA, cross-ancestry meta-analyses were used, whereas CanUD PGS incorporated population-concordant and ancestry-specific SNPs (**Supplemental Table 16**).

Sex-stratified analyses are presented in **Figure 3** and **Supplemental Table 17**. Sample sizes for CanUD and OUD in IB were insufficient for sex-stratified analyses. Seven female and six male PGS did not reach statistical significance, with 10 from AA and three from LA. Overall, male PGS generally showed higher or comparable ORs relative to female PGS, except for AUD and polySUD in AOU LA and TUD in IB LA. Notably, female AUD PGS in AOU LA showed significantly higher ORs than male AUD PGS, whereas all other male-female comparisons had overlapping 95% CIs.

**Figure 3:**
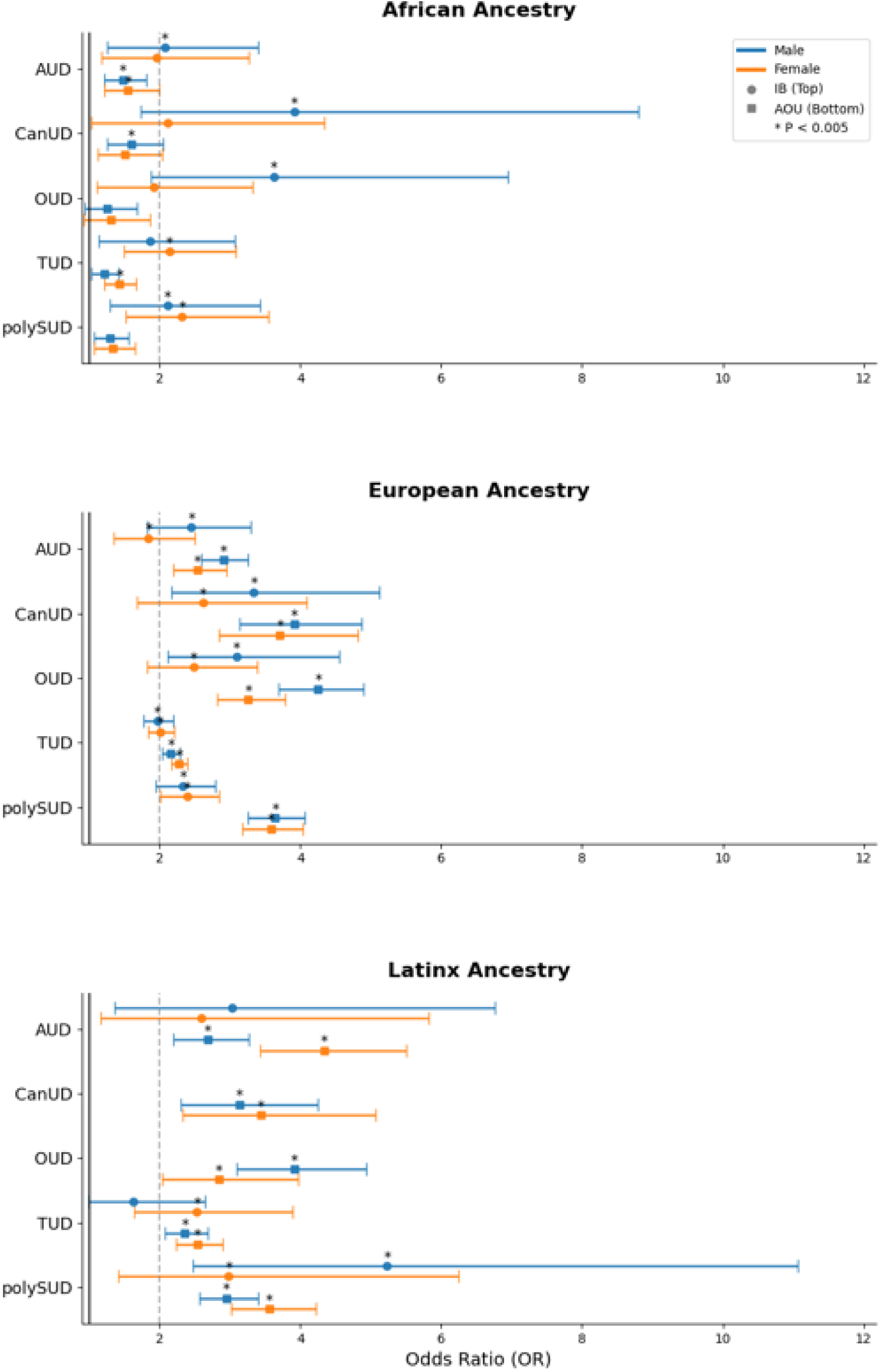
Sex-stratified analysis results: Note: AUD: alcohol use disorder; CanUD: cannabis use disorder; OUD: opioid use disorder; TUD: tobacco use disorder; polySUD: poly substance use disorder; Dashed line indicates OR=2.

## DISCUSSION

In this study, we applied the two-stage optimization and validation framework to develop and validate PGS for AUD, CanUD, OUD, TUD, and polySUD across AA, EA, and LA. In both IB and AOU, all PGS in EA and LA achieved the predefined clinically relevant threshold (OR ≥2), with approximately half having ORs ≥3. Notably, several PGS demonstrated large effect sizes, particularly for OUD, CanUD, and polySUD in LA, although 95% CIs were wider due to small sample sizes. In contrast, although most PGS in AA showed statistically significant associations, predictive performance did not meet the clinically relevant threshold in AOU. Sex differences in PGS performance were generally modest.

Selection of discovery datasets and PGS methods substantially influenced predictive performance. Unexpectedly, with few exceptions, the best-performing PGS were derived from crossSUD GWAS rather than disorder-specific GWAS. Although crossSUD GWAS did not capture disorder-specific SNPs, they were based on substantially larger sample sizes, likely enabling more accurate estimation of effect sizes and improved identification of SUD-shared SNPs. These findings suggest that shared genetic liability across SUDs may contribute more strongly to prediction than disorder-specific SNPs. In AA and LA populations, all best-performing discovery datasets were based on cross-ancestry meta-analyses, indicating that many SUD-associated SNPs are shared, and confirming that leveraging large-scale EA GWAS can improve predictive performance in underrepresented populations(30). Incorporation of population-concordant and ancestry-specific SNPs improved predictive performance only for CanUD PGS, possibly because currently available ancestry-specific GWAS remain underpowered relative to EA GWAS. PGS performance was substantially lower in AA than in LA, potentially reflecting the larger proportion of EA ancestry in LA(39). We also observed that no single PGS construction method consistently outperformed others. Approximately half of the best-performing PGS were generated using P+T, whereas the remainder used PRS-CS or PRS-CSx, underscoring the importance of systematically evaluating multiple approaches using the two-stage framework when optimizing PGS performance.

Generalizability remains a major challenge for clinical implementation of PGS. PGS developed in one cohort frequently showed reduced performance in independent datasets, particularly when ascertainment strategies differ substantially. To improve generalizability, we intentionally did not use cohorts specifically enriched for SUDs, despite their potentially greater statistical power. Instead, we used two large biobank-based cohorts with SUD prevalences closer to those observed in the general U.S. populations. Because IB represents a regional health system cohort whereas AOU is a national cohort with participants from diverse backgrounds and health conditions, validation in AOU provided a stringent test of reproducibility and generalizability.

We required that the high-risk thresholds remain below the prevalence of the corresponding disorder; however, among PGS meeting criteria for clinical relevance, those at high-risk were not limited to the extreme upper tail of the PGS distribution (i.e., only identifying a small number of high-risk individuals). Most thresholds exceeded 50% of the disorder prevalences, and approximately one-third approached 90% of the prevalences, suggesting that clinically meaningful risk prediction may be achievable in a relatively large proportion of the population. Although SUD prevalence was generally higher in males than in females, sex differences in PGS performance were modest overall. Notably, in AOU LA, males had substantially higher AUD prevalence than females, whereas female AUD PGS demonstrated significantly higher predictive performance. These findings suggested that sex differences in SUD prevalences may be influenced more strongly by environmental factors than by differences in genetic liability alone.

Although these findings support the potential clinical implementation of PGS in predicting SUD risks, several important considerations should be emphasized. First, PGS are not intended for diagnosis or prognosis, which should rely on clinical and symptom-based assessments. Second, individuals with high PGS are not destined to develop SUDs, environmental exposures and social factors strongly influence disease onset. Accordingly, PGS should be interpreted cautiously and should never be used to discriminate against individuals or restrict access to insurance, prevention, or treatment services. Finally, reduced predictive performance in AA populations should not be interpreted as evidence of distinct underlying biological mechanisms. Rather, these findings underscore the urgent need for larger and more diverse genetic studies to reduce disparities in genomic medicine(40).

This study has several limitations. First, the prevalences of SUDs in IB and AOU were not identical to those in the general U.S. populations; therefore, the generalizability of these PGS requires further evaluation. Second, we evaluated only three PGS methods and did not incorporate functional annotation of SNPs; thus, alternative approaches may further improve PGS predictive performance. Third, LA sample size was relatively small, limiting our ability to conduct well-powered sex-stratified analyses. In addition, insufficient sample sizes precluded PGS analyses in populations beyond AA, EA, and LA. Fourth, case and controls were identified using ICD codes rather than structured diagnostic interviews. Misclassification may have occurred, particularly among controls who may have undiagnosed SUDs.

In summary, we developed and validated PGS for the five most prevalent SUDs using the two-stage optimization and validation framework. Beyond demonstrating clinically informative risk stratification in EA and LA populations, our findings support the application of this framework for developing robust and generalizable PGS across diverse populations. Future work will focus on improving predictive performance in AA and extending PGS development to additional ancestral populations. In addition, because the prevalences of SUDs in IB and AOU are not identical to those of the general U.S. populations, further efforts are needed to enhance generalizability, including the use of resampling approaches to better approximate population-representative cohorts.

## Data Availability

All data produced in the present work are contained in the manuscript or will be available at PGS catalog.

## ACKNOWLEDGMENTS

The All of Us Research Program is supported by the National Institutes of Health, Office of the Director: Regional Medical Centers: 1 OT2 OD026549; 1 OT2 OD026554; 1 OT2 OD026557; 1 OT2 OD026556; 1 OT2 OD026550; 1 OT2 OD 026552; 1 OT2 OD026553; 1 OT2 OD026548; 1 OT2 OD026551; 1 OT2 OD026555; IAA #: AOD 16037; Federally Qualified Health Centers: HHSN 263201600085U; Data and Research Center: 5 U2C OD023196; Biobank: 1 U24 OD023121; The Participant Center: U24 OD023176; Participant Technology Systems Center: 1 U24 OD023163; Communications and Engagement: 3 OT2 OD023205; 3 OT2 OD023206; and Community Partners: 1 OT2 OD025277; 3 OT2 OD025315; 1 OT2 OD025337; 1 OT2 OD025276. In addition, the All of Us Research Program would not be possible without the partnership of its participants.

This study was made possible, in part, with support from the Indiana Clinical and Translational Sciences Institute funded, in part by Award Number UL1TR002529 from the National Institutes of Health, National Center for Advancing Translational Sciences, Clinical and Translational Sciences Award, and the National Center for Research Resources, Construction grant number RR020128 and the Lilly Endowment. The content is solely the responsibility of the authors and does not necessarily represent the official views of the National Institutes of Health.

The authors acknowledge the Indiana University Pervasive Technology Institute for providing [HPC (Big Red II, Karst, Carbonate), visualization, database, storage, or consulting] resources that have contributed to the research results reported within this paper.

D. Lai and Y. Liu are supported by National Institutes of Health award number AA031176.

