## Supplemental Methods for "Toward Clinical Implementation of Polygenic Scores for Substance Use Disorders: A Multi-Ancestry Study"

*Data processing:*

Indiana Biobank (IB)(1, 2) data was whole exome sequenced at 30X coverage and targeted low-pass whole genome sequenced at 2X coverage, then imputed using GLIMPSE2.0 (Genotype likelihoods Imputation and phasing method)(3) with 1000 Genomes Project(4) data as the reference panel. Variants with imputation quality score (INFO score) ≥0.8 were retained. Imputed allele dosages were converted to hard-call genotypes using the following thresholds: dosages <0.1 were assigned as homozygous for the non-reference allele, between 0.9 and 1.1 as heterozygous, >1.9 as homozygous for the reference allele, and those not meeting these thresholds were set as missing. Then variants with genotyping rate <95%, minor allele frequency <1%, Hardy-Weinberg equilibrium P-value <5.0E-7 (for autosomal variants) were excluded. Principal components (PCs) of genetic ancestry were calculated using SNPRelate(5) with 1000 Genomes Project(4) samples as the reference data. A small portion of IB samples were related, and a set of unrelated samples were selected based on disease status and genotyping rates.

All of Us Research Program (AOU, version 8)(6) data was whole genome sequenced. Due to the data use policy, all data processing and analyses were performed at the AOU workbench. We used PLINK(7, 8) datasets that have population-specific allele frequency >1% or population-specific allele count >100 in any ancestry subpopulation provided by AOU. Similar to IB, we selected a set of unrelated samples based on disease status and genotyping rates.

For both IB and AOU, samples with mismatched genetically determined and reported sex were removed; and only genetically determined African (AA), European (EA), and Latinx (LA) samples based on PCs were included in the analysis. For all datasets (including GWAS summary statistics), palindromic variants (e.g., A/T or C/G variants) were excluded due to the strand ambiguity. Only those variants with rs numbers and passed QC in all datasets were retained.

*Definition of cases and controls in target datasets:*

In IB, SUD case-control status was determined using ICD-9/10 codes. The goal of PGS analyses is to maximize identification of individuals at high-risk. Therefore, we used a broader definition, and individuals having at least one related ICD code were considered as cases to reduce potential false negatives among high-risk individuals. The following ICD codes were used: 303 and 305.0 (ICD9), F10 (ICD10) for AUD; 304.3 and 305.2 (ICD9), F12 (ICD) for CanUD; 304.0 and 305.5 (ICD9), F11 (ICD10) for OUD, 305.1 (ICD9), F17 (ICD10) for TUD. For each SUD, individuals that did not have related ICD codes were considered as controls (i.e., they may have other SUDs). For polySUD, we also used codes for cocaine use disorder (ICD9: 304.2 and 305.6; ICD10: F14) and other SUDs (ICD9: 304.1, 304.4, 304.5, 304.6, 304.7, 304.8, 304.9, 305.3, 305.4, 305.7, 305.8, and 305.9; ICD10: F13, F15, F16, F17, F18, and F19). Individuals having at least two different SUDs were considered as polySUD cases and all others were considered as controls.

In AOU, case-control status was determined using ICD-9/10 codes mapped through the OMOP common data model(6). The following OMOP concept IDs were used: 4218106 (AUD), 440387 and 434327 (CanUD), 438120 and 438130 (OUD), 4209423 (TUD). For each SUD, individuals that did not have corresponding OMOP concept IDs were considered as controls. For polySUD, the following OMOP concept IDs were also used: 432303 and 436389 (cocaine use disorder), 40479573 (stimulant abuse), 443236 (hypnotic or anxiolytic dependence), 442601 (sedative abuse). Individuals having at least two different SUDs were considered as polySUD cases and all others were considered as controls.

All cases and controls were ≥18 years old except AOU controls, which were ≥21 years old.
